# The Trends of Physical Morphology, Overweight, and Obesity of The Elderly in Xinjiang From 2000 to 2030

**DOI:** 10.1101/2025.08.01.25332575

**Authors:** Yuanyuan Ding, Chunjing Tu, Xinzhe Pu

## Abstract

This study aims to investigate the changes and trends in body shape, overweight, and obesity among the elderly in the Xinjiang Uyghur Autonomous Region from 2000 to 2020. Additionally, it seeks to predict these trends for the next 10 years, providing valuable insights for enhancing the health of the elderly in Xinjiang. Based on the body morphology data of the elderly collected from physical fitness monitoring in Xinjiang in 2000, 2005, 2010, 2014, and 2020, a grey GM(1,1) model was established to explore the dynamic characteristics and future trends of each index. The results showed that all physical indicators for the elderly in Xinjiang showed an upward trend between 2000 and 2020, During this period, the rates of overweight, obesity, and central obesity among the elderly also continued to rise. Specifically, the increase in rates for males was 3.1% for overweight, 13.4% for obesity, and 27.4% for central obesity. For females, the increases were 3.5%, 13.7%, and 21.4%, respectively. These trends are expected to continue over the next decade, with all indicators likely to keep increasing, particularly as central obesity rates rise faster than general obesity rates. In conclusion, the rates of overweight and obesity among the elderly in Xinjiang are increasing year by year. Measures should be taken at multiple levels, including policy, society, and individuals, to curb the prevalence of overweight/obesity, thereby promoting the physical health of the elderly in Xinjiang.

## Introduction

Body shape indicators can well judge health status, especially if the weight level is very close to health. Currently, the problem of obesity among the elderly in China is becoming serious, and the World Obesity Blueprint points out that obesity in China is currently high in absolute terms and rising [1] and studies have shown that obesity increases the risk of developing type 2 diabetes, cardiovascular, and hypertensive chronic diseases, and also negatively affects mental health and socio-economics,[2,3] and not only affects the progress of the “Healthy Aging Plan”, but also hurts the high-quality development of the population. This not only affects the progress of the Healthy Aging Plan but also hurts the quality development of the population.[4] In 2024, 16 Chinese governmental departments will jointly implement the “Year of Weight Management”, which calls for the strengthening of weight management for the elderly.[5] Xinjiang has also introduced the Xinjiang Uygur Autonomous Region’s “14th Five-Year Plan for the Development of the Elderly and the Elderly Service System” to protect the health of the local elderly.[6] Xinjiang Uygur Autonomous Region is located on the northwestern border of China, with a large number of ethnic groups, and a unique natural and social environment that creates physical characteristics that are different from those of other provinces and cities.[7]

Based on this, this study used the data on body morphology and other data of the elderly obtained from five times of physical fitness monitoring in Xinjiang from 2000 to 2020, and indexed the establishment of a gray GM (1,1) prediction model of the series of indicators of body morphology of the elderly in Xinjiang, to study the development trend of the body morphology and the development of the development trend of overweight and obesity in the elderly in Xinjiang, and to provide a reference for the improvement of the health of elderly people in Xinjiang.

## Methods

### Study population and data source

The study’s object was the body morphology of elderly people in Xinjiang from 2000 to 2020. We extracted data from the series of data obtained from five national physical fitness monitoring sessions in 2000, 2005, 2010, 2014, and 2020, and the principle of randomized cluster sampling was adopted for the selection of samples at all times.

### Criteria for Determining Indicators

The main indicators needed for body shape included height, weight, waist circumference(WC), and body mass index (BMI). The Chinese obesity BMI standard defined in (WS/T428-2013) (24kg/m^2^ ≤ BMI ≤ 27.9kg/m^2^ for overweight, BMI ≥ 28kg/m^2^ for obesity) and the central obesity standard (≥ 85cm waist circumference for women and ≥ 90cm waist circumference for men for central obesity) were applied.[8]

### Research method

The main research methodology of this study is to construct a gray GM (1, 1) forecasting model using the successive monitoring data as time series data, fit the curves of growth and development indicators, and then analyze the dynamics of the 20 years from 2000 to 2020.

## Results

### Trends and predicted results of body shape changes in elderly people from 2000 to 2030 in Xinjiang Uygur Autonomous Region

According to the methods and steps of gray mean GM (1, 1) modeling, the trend model of physical morphology indicators of the elderly in Xinjiang district was constructed. The parameters, fitted values, and predicted values could be derived, among which, the parameter a represents the model development coefficient, which reflects the intrinsic trend of the time-series data, with a “negative” indicating the increasing trend of the series, and an A “negative” a indicates an increasing trend in the series, a “positive” indicates a decreasing trend in the series, and the larger the absolute value of a is, the larger the magnitude of increase or decrease is, and vice versa. Commonly used indicators of body shape are mainly height, weight, waist circumference, and derived indicators such as BMI, etc. Through the gray GM (1,1) model, it can be seen that the development coefficient of all the indicators of the two age groups male and female, are all negative (**Error! Reference source not found**.), which indicates that all the indicators of the body shape have been in an increasing trend since 2000, and the specific analyses are as follows.

### Height

Between 2000 and 2020, the height of elderly males increased by 1.45 cm and the height of elderly females increased by 1.7 cm, representing growth rates of 0.17% and 0.29% every five years, respectively. It is projected that from 2020 to 2030, the height of elderly males and females will rise by 0.18% and 0.27% every five years, respectively. Over the next decade, the height of the elderly population in China is expected to continue to grow, although at a slightly lower rate compared to the prior two decades(TABLE 1, FIG 1.A).

**TABLE 1.**
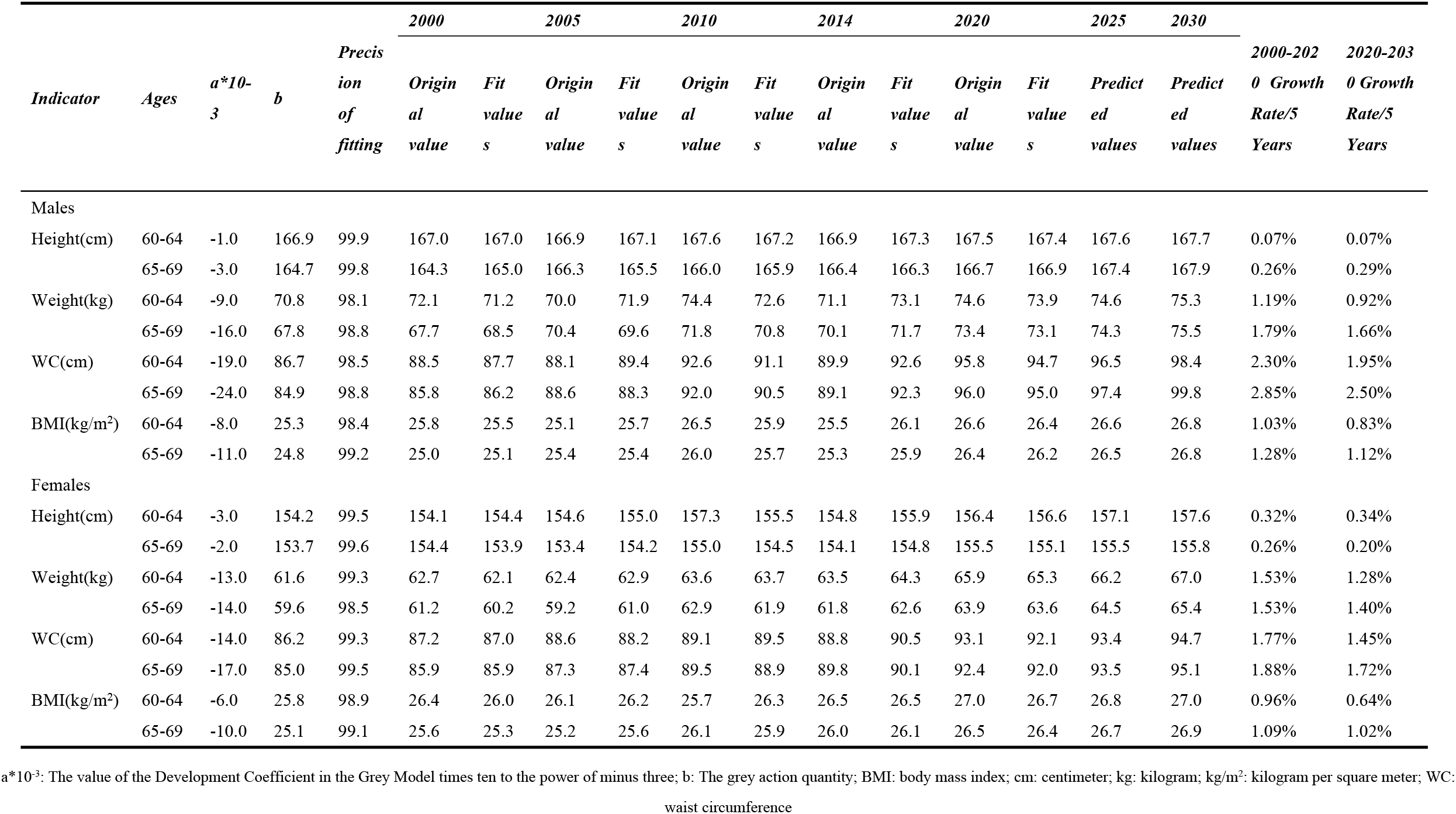
List of Trends and Gray Projections of Changes in Body Morphology of the Elderly in Xinjiang Uygur Autonomous Region, 2000-2030.

**FIG 1.**
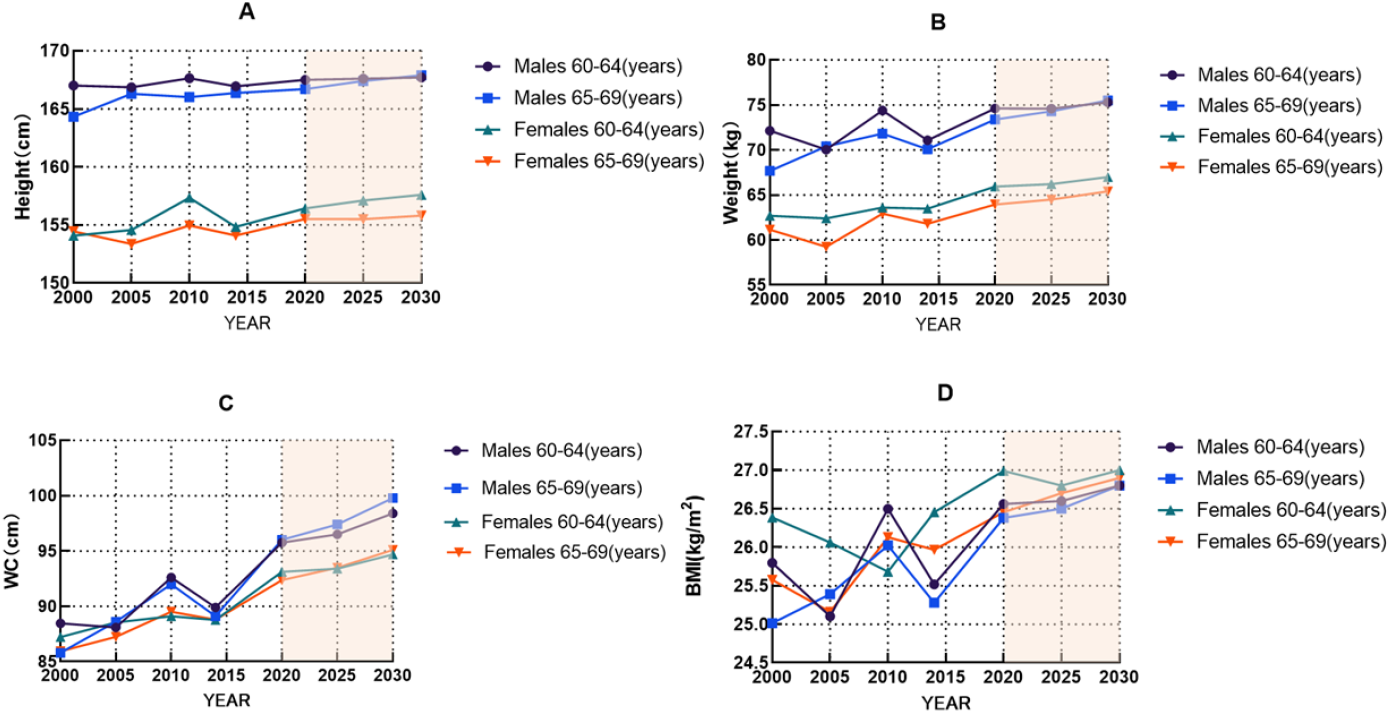
Changes in mean values and projections of height (A), weight (B), waist circumference (C), and BMI (D) indicators for the elderly in Xinjiang District, 2000-2030. BMI:body mass index; cm: centimeter; kg/m2: kilogram per square meter; WC: waist circumference

### Weight

Between 2000 and 2020, the body weight of elderly males increased by 3.7 kg, while elderly females saw an increase of 3.3 kg. This corresponds to a growth rate of 1.49% per five-year period for males and 1.53% for females. Projections indicate that from 2020 to 2030, the body weight of elderly males and females will rise by 1.29% and 1.34% per five-year period, respectively. Over the next ten years, the weight of the elderly population in China is expected to continue increasing, albeit at a slightly lower rate compared to the prior two decades(TABLE 1, FIG 1.B).

### Waist circumference

Between 2000 and 2020, the WC of older adults increased by 7.9 cm for males and 5.6 cm for females, which represents growth rates of 2.57% and 1.83% every five years, respectively. Projections for the period from 2020 to 2030 suggest that the waist circumference of males and females will increase at rates of 0.18% and 0.27% every five years, respectively. Over the next decade, the waist circumference of the elderly in China is expected to continue increasing, although at a slightly lower rate than observed in the prior two decades(TABLE 1, FIG 1.C).

### BMI

Between 2000 and 2020, the BMI of older males increased by 1.0, while older females experienced an increase of 0.9. This corresponds to an increase of 1.16% for males and 1.03% for females every five years. It is projected that from 2020 to 2030, the BMI of older males and females will increase by 0.98% and 0.83% per five-year period, respectively. Over the next decade, the BMI of the elderly in China is expected to continue to rise, although at a slightly slower rate compared to the prior two decades(TABLE 1, FIG 1.D)

### Trends in the detection rates of overweight, obesity, and central obesity among the elderly in Xinjiang district

The Chinese obesity BMI standard (24 ≤ BMI ≤ 27.9 for overweight, BMI ≥ 28 for obesity) and the central obesity standard (≥ 85 waist circumference for females and ≥ 90 waist circumference for males for central obesity) were used to identify the data from the physical fitness survey of the elderly in Xinjiang Uygur Autonomous Region. The results were found (see TABLE 2):(1) In terms of chronological changes. Firstly, the overweight rate, obesity rate, and central obesity rate of men increased by 3.1%, 13.4%, and 27.4% respectively between 2000 and 2020, while the increase of these three values of women was 3.5%, 13.7%, and 21.4% respectively, and the characteristics of the increase were similar to those of men, with the growth of overweight, obesity as well as central obesity, and the central obesity rate has developed most rapidly; the forecast found that in the next 10 years, the overweight rate, obesity rate, and central obesity rate will still maintain rapid growth, with a predicted increase of 1.3%, 11.8%, and 19.7% for males and 1.7%, 12.2%, and 16.5% for females in 10 years. Secondly, from 2000 to 2020, the overweight rate, obesity rate, and central obesity rate in Xinjiang will increase by 2.9%, 27.9%, and 22.2% for males and 3.8%, 31.1%, and 18.4% for females respectively every five years, and both of them will have the highest growth rate of obesity rate; from 2020 to 2030, it is predicted that the growth will still be maintained, with overweight rate, obesity rate, and central obesity rate growing at 2.2%, 23.2%, and 16.9% per 5 years for males, and 3.2%, 24.7%, and 16.3% for females, respectively, which is a decrease in growth rate compared to the previous two decades.

**TABLE 2.** Detection rates of overweight, obesity, and central obesity among the elderly in Xinjiang, 2000-2030 (%)

| *Project* | *2000* | *2005* | *2010* | *2014* | *2020* | *2025* | *2030* | *2000—2020* | *2020—2030* |
| --- | --- | --- | --- | --- | --- | --- | --- | --- | --- |
|  |  |  |  |  |  |  |  | *Growth Rate/5 Years* | *Growth Rate/5 Years* |
| Overweight rate of Males | 26.8 | 27.2 | 33.0 | 28.1 | 29.9 | 30.5 | 31.2 | 2.9 | 2.2 |
| Overweight rate of females | 23.0 | 24.5 | 24.9 | 25.4 | 26.5 | 27.4 | 28.2 | 3.8 | 3.2 |
| Obesity Rate of Males | 12.0 | 15.4 | 13.9 | 20.8 | 25.4 | 30.8 | 37.2 | 27.9 | 23.2 |
| The obesity rate of females | 11.0 | 14.5 | 13.5 | 19.8 | 24.7 | 30.7 | 36.9 | 31.1 | 24.7 |
| The central obesity rate of Males | 30.9 | 38.2 | 44.7 | 48.4 | 58.3 | 67.4 | 78.0 | 22.2 | 16.9 |
| The central obesity rate of females | 29.1 | 34.0 | 34.3 | 43.4 | 50.5 | 58.2 | 67.0 | 18.4 | 16.3 |

Note: 2025 and 2030 are projections from the gray GM(1,1) model, and national data are shown in previous National Physical Fitness Monitoring Survey bulletins or reports.

## Discussion

We extracted data from five large-scale physical fitness surveys in Xinjiang District since 2000 to provide a comprehensive overview of long-term trends in height, weight, waist circumference, and BMI covering more than 25 million people in Xinjiang District, and we analyze the relevant findings as follows.

### Chronological trends and causes of changes in body shape indicators of the elderly in the Xinjiang region, 2000-2030

This study found that from 2000 to 2020, all age groups of body morphology indicators of the elderly in Xinjiang district were on the trend of growth. The growth rate of waist circumference > weight > height, which is consistent with the trend of development of BMI, and the indicators of body morphology will continue to increase from 2020 to 2030. This is consistent with the trend of chronological changes in body morphology of the elderly in cities and towns in China, as well as the trend of changes in body morphology of the local elderly in Karamay City, Xinjiang;[9,10] the growth trend of the chronological changes in height of the elderly in Xinjiang is slow, which is similar to the trend of the chronological changes in the elderly in the whole country.[11]

The reasons for the changes in physical indicators come from a number of sources, starting with the significant economic development, the emergence of over-nutrition, and the increase in social security. In addition, the lack of a qualitative increase in the level of exercise and changes in lifestyle may also be a factor.

### Characteristics and Causes of Changes in Overweight and Obesity among the Elderly in Xinjiang Region, 2000-2030

From 2000 to 2020, the overweight and obesity rates of the elderly in Xinjiang showed a rapidly increasing trend, with an increase of 3.3% and 13.55% respectively, and will continue to increase in the next 10 years. In the same period, the overall change of overweight/obesity of the elderly in China also shows an increasing trend, some studies have shown that from 2000-2014 the overweight/obesity rate of the elderly in China increased by 6.9% and 3.1%,[11] and in the Fifth Physical Fitness Monitoring Bulletin for the Entire Population, in 2014-2020, overweight/obesity of the elderly increased by 0.1% and 2.5%, respectively. Increased by 0.1% and 2.8%,[12] in which the increase of obesity rate in Xinjiang was higher than that of the whole country, which may be due to the unique dietary habits and humanistic characteristics of Xinjiang region. In addition, the tracking study of healthy longevity in China’s elderly also pointed out that the overweight/obesity rate of the nation’s elderly increased year by year from 2008 to 2018.[13] The main reasons for the continuous increase in overweight/obesity rates are as follows.

First, Economic factors: The level of economic development is one of the key factors affecting national fitness. Xinjiang’s GDP was 136.356 billion yuan in 2000, while in 2020 it rose to 137.758 billion yuan, a tenfold increase, while the per capita GDP went from 7,372 yuan to 53,593 yuan, a 7.27-fold increase. Some studies have shown a positive correlation between GDP per capita and overweight and obesity,[14] as GDP per capita grows in Xinjiang, the quality of the diet improves, but it is prone to over-nutrition, and the overweight phenomenon becomes more serious.[15]

Second, Social security factors: The increase in income brought about by economic growth has led to an increase in the level of social security benefits,[16] and the increase in the level of social security will lead to an increase in the prevalence of overweight/obesity,[17] and the number of people enjoying pension benefits in Xinjiang District has increased from 1,057,900 in 2015 to 1,592,600 in 2023, and the proportion of elderly people receiving pensions is getting higher,[18,19] and with the current proportion of in-service participation reaching more than 90%, it is expected that the proportion of elderly people receiving pensions will be even higher in the future, so overweight/obesity tends to be serious.

Third, Physical exercise factor: Physical activity is a protective factor affecting overweight/obesity in older adults, and a decrease in physical activity increases the trend toward overweight/obesity.[20] In the medical examination of a division city of Xinjiang Corps in 2018-2023, it was found that 60.03% of the elderly people in 2023 did not participate in exercise increased by 1.57 percentage points compared to 2018, while those who exercised every day decreased by 3.3% percentage points.[21] Despite the improvement in physical activity among older people in Xinjiang noted in several studies,[22] the overall results are poor, and older people in Xinjiang still face a lack of exercise.

Fourth, Ethnic factors: In 2020, the resident population of Xinjiang Uygur Autonomous Region accounted for 42.24% of Han Chinese, 44.96% of the Uygur population, and 12.80% of other ethnic minorities.[23] The region has a large number of ethnic groups, with large mixed populations and small clusters, and most studies have shown that obesity among most of Xinjiang’s ethnic minorities is higher than that of the Han Chinese,[12,24] with the Uyghur population growing from 8,345,600 in 2000 to 11,624,300 by 2020, at an average annual growth rate of 1.67%, much higher than the national average growth rate of ethnic minority populations of 0.83% during the same period. The white paper “Population Development in Xinjiang” noted that there are more women of childbearing age among ethnic minorities in Xinjiang that the future growth of the ethnic minority population has great potential,[25] and that the problem of overweight/obesity will further increase in the future.

Fifth, Lifestyle factors: With the rapid development of the urbanization level in Xinjiang District, the proportion of the urban resident population has grown from 43.01% in 2010 to 57.89% in 2022,[26] the rising level of urbanization has brought about changes in lifestyles, and people’s dietary structure and lifestyles may change in the process of urbanization For example, people have more social functions and will consume more high-fat food, use private cars for commuting and more sedentary behaviors are all reasons for higher obesity rates.

Comparing central obesity with generalized obesity among the elderly in the Xinjiang region. The prevalence of central obesity in the elderly in Xinjiang district during 2000-2020 was much higher than the prevalence of generalized obesity based on BMI. This phenomenon is similar to the changes in body shape of urban adults in China.[27] This may be related to the distribution of body composition in Asian populations, i.e., at the same BMI, there will be more fat and more distributed in the abdomen.[27,28] Sun Jieying’s study found that participation in physical exercise was negatively correlated with the growth of waist circumference, while sedentary behavior showed a positive correlation with the growth of waist circumference,[29] on the one hand, the ideological awareness of exercise among the elderly in Xinjiang is weak, the exercise venues are insufficient, and the organization of the jagged leads to insufficient exercise in the elderly [21] making overweight/obesity risk increase; on the other hand, with the change of lifestyle in Xinjiang Xinjiang Increase in sedentary time among older adults leads to obesity. Intake of high-calorie and high-fat food is prone to central obesity,[30] meat consumption in Xinjiang has risen from 23.2g per capita in 2015 to 26.9g per capita in 2022 and shows an upward trend and high-calorie and high-fat food has continued to show an upward trend in recent years, which is expected to continue to increase in the future, and it is expected that central obesity rates will continue to be higher than generalized obesity rates in the future.

## Conclusion

In general, the body shape, overweight, obesity, and central obesity of the elderly in Xinjiang Region showed an increasing trend from 2000 to 2020; it is predicted that each indicator will continue to grow in the next 10 years.

The level of economic development, regional customs, physical exercise behavior, and lifestyle changes are important factors affecting the physical form, overweight, and obesity of the elderly in Xinjiang, and government departments at all levels should adopt more effective intervention policies and measures to curb the trend of weight gain and improve the physical health of the population.

## Data Availability

The data underlying the results presented in the study are available from corresponding author upon reasonable request.

## Acknowledgments

We are sincerely thankful to the Xinjiang Uyghur Autonomous Region Sports Science Research Center and all subjects being test volunteers and testers for their assistance in our study.

